# Years of Life Lost in the United States During the COVID-19 Pandemic, March 2020 to October 2021

**DOI:** 10.1101/2022.07.22.22277899

**Authors:** Mark É. Czeisler, Charles A. Czeisler

## Abstract

**Background:** Given a downward age shift in COVID-19-involved deaths observed during the COVID-19 pandemic, we sought to estimate years of life lost (YLL) associated with leading causes of US death during the first 20 months of the pandemic.

**Findings:** Despite 4796 fewer COVID-19 deaths in Jan-Oct 2021 than in Mar-Dec 2020, the number of YLL due to COVID-19 increased by 1,159,761, from 4,474,186 to 5,633,947 (a 25.9% increase). YLL per COVID-19 death increased from 12.8 in 2020 to 16.3 in 2021, a 27.7% increase. YLL per death did not change by more than 2.3% for any other cause.

**Interpretation:** Increased YLL per COVID-19 death in 2021 result from younger-age COVID-19 mortality, contributing to a marked increase in YLL from this preventable cause of death at a later stage of the pandemic despite advancements in vaccines in treatments.

## 1. Introduction

More than a million excess deaths have occurred in the United States during the COVID-19 pandemic.^1^ As recently described by Shiels and colleagues in *JAMA Internal Medicine*,^2^ COVID-19 deaths during March through December 2020 and January through October 2021 were similar. However, COVID-19-involved deaths increased among younger persons and decreased among older adults in 2021 versus 2020,^2^ reflecting excess premature mortality from COVID-19. Quantifying the magnitude of this downward age shift in COVID-19-involved deaths requires an age-weighted metric. Unlike the mortality metric, potential years of life lost (YLL)^3^ offers an indicator of premature mortality based on the estimated number of years a person would have lived if they had not died prematurely. We therefore sought to estimate YLL associated with leading causes of US death during the first 20 months of the pandemic.

## 2. Methods

### 2.1. Data Sources

March 2020 through October 2021 mortality data by age and sex were obtained from the National Center for Health Statistics.^4^ The 15 leading causes of US deaths occurring between March and December 2020 and between January and October 2021 were identified. The causes were the same, with some variations in order. For each study interval, deaths due to each of the 15 leading causes were downloaded by sex and age. Deaths during the study intervals that had insufficient information on sex and age (either due to an age listed as “Not Stated” or to a Suppressed death count, which occurs when the data meet the criteria for confidentiality constraints) were excluded from the analysis.

Leading causes of death and associated ICD codes follow: heart disease (I00-I09, I11, I13, I20-I51); cancer (C00-C97); COVID-19 (U07.1); unintentional injuries (V01-X59, Y85-Y86); stroke or cerebrovascular diseases (I60-I69); chronic lower respiratory diseases (J40-J47); Alzheimer disease (G30); diabetes mellitus (E10-E14); chronic liver disease and cirrhosis (K70, K73-K74); nephritis, nephrotic syndrome and nephrosis (N00-N07, N17-N19, N25-N27); influenza and pneumonia (J09-J18); intentional self-harm or suicide (*U03, X60-X84, Y87.0); essential hypertension and hypertensive renal disease *I10, I12, I15); Parkinson disease (G20-G21); and septicemia (A40-A41).

The most recent (2019) standard life expectancies (SLEs) by age and sex were obtained from the National Vital Statistics System through period life tables.^5^ Period life tables provide estimates of how many more years a group of people who are currently at a particular age – any age from birth to 100 years or more – can expect to live if the mortality patterns in a given year remain the same over the rest of their lives. Standard life expectancy estimates were downloaded by age and sex given differences by these characteristics. Although life expectancy differs by combined race and ethnicity,^6^ these characteristics were intentionally excluded from estimates of years of life lost given the nature of race as a social construct and the systemic devaluing of some lives on the basis of race and ethnicity that would occur if included.

### 2.2. Statistical Analysis

For each cause of death, total years of life lost were estimated by multiplying the number of deaths in each single-age group for each sex by the SLE of that demographic. For example, there were 95 heart disease deaths among 25-year-old men, who had a SLE of 52.5 years, corresponding to 4,987.5 YLL. Among 25-year-old women, who had a SLE of 57.2 years, there were 54 heart disease deaths, corresponding to 3,088.8 YLL. YLL were totaled across all ages and both sexes for each cause of death. YLL were estimated for the 15 leading causes of death, comparing March through December 2020 with January through October 2021. YLL per death were also estimated for each interval by dividing the number of years of life lost by the number of deaths. Data were accessed on July 7, 2022, and analyzed in Python version 3.7.8. This study used publicly available data and was not subject to institutional review.

## 3. Results

The 15 leading causes of US death were the same in both 10-month intervals and accounted for 81% of deaths in each. Unsuppressed age-and sex-characterized data were available for 99.94% of deaths in each interval.

In 2020 and 2021, the 15 leading causes of deaths were associated with 34,387,967 and 35,944,895 potential years of life lost, respectively. The number of YLL for the two leading causes of YLL were stable and consistent across intervals: cancer (2020, 7,718,240 YLL; 2021, 7,710,720 YLL) and heart disease (2020, 7,204,329 YLL; 2021, 7,202,137 YLL) (Table). For the third-leading cause of YLL, unintentional injuries, YLL increased by 8.5% (2020, 5,268,068 YLL; 2021, 5,714,413 YLL), comparable to the 8.7% increase in deaths from unintentional injuries. In contrast, despite 4796 fewer COVID-19 deaths in 2021 than in 2020, the number of YLL due to COVID-19 increased by 1,159,761, from 4,474,186 to 5,633,947 (a 25.9% increase).

**Table.** Leading Causes of US Death and Associated Potential Years of Life Lost, March to December 2020 and January to October 2021.

| Cause* | Deaths |  | Years of Life Lost |  | Years of Life Lost per Death |  |
| --- | --- | --- | --- | --- | --- | --- |
|  | March to December 2020 | January to October 2021 | March to December 2020 | January to October 2021 | March to December 2020 | January to October 2021 |
|  | N | N | N | N | N | N |
| Heart disease | 579,424 | 571,228 | 7,204,329 | 7,202,137 | 12.4 | 12.6 |
| Cancer | 501,105 | 502,489 | 7,718,240 | 7,710,720 | 15.4 | 15.3 |
| COVID-19 | 350,702 | 345,906 | 4,474,186 | 5,633,947 | 12.8 | 16.3 |
| Unintentional injuries** | 171,626 | 186,607 | 5,268,068 | 5,714,413 | 30.7 | 30.6 |
| Stroke | 133,227 | 133,788 | 1,463,097 | 1,493,362 | 11.0 | 11.2 |
| Chronic lower respiratory diseases | 122,686 | 116,255 | 1,521,205 | 1,446,674 | 12.4 | 12.4 |
| Alzheimer disease | 112,613 | 98,104 | 760,801 | 673,835 | 6.8 | 6.9 |
| Diabetes | 86,111 | 84,902 | 1,355,279 | 1,350,773 | 15.7 | 15.9 |
| Chronic liver disease and cirrhosis | 43,784 | 46,802 | 1,034,197 | 1,113,108 | 23.6 | 23.8 |
| Kidney disease | 43,041 | 44,533 | 545,716 | 574,354 | 12.7 | 12.9 |
| Influenza and pneumonia | 40,599 | 33,305 | 512,907 | 424,624 | 12.6 | 12.7 |
| Suicide | 38,444 | 40,224 | 1,339,733 | 1,409,995 | 34.8 | 35.1 |
| Hypertension | 35,102 | 34,925 | 409,681 | 416,954 | 11.7 | 11.9 |
| Parkinson disease | 33,762 | 31,299 | 290,242 | 272,978 | 8.6 | 8.7 |
| Septicemia | 32,897 | 33,723 | 490,288 | 507,021 | 14.9 | 15.0 |
| **15-cause total** | **2,325,123** | **2,304,090** | **34,387,967** | **35,944,895** | **-** | **-** |

\* Causes of death are listed by leading causes of death in 2020.
\*\* Unintentional injury deaths include transportation incidents, unintentional drug overdoses and alcohol-related deaths, and other unintentional injury deaths.

COVID-19 was distinct with regards to changes to the number of YLL per death. YLL per COVID-19 death increased from 12.8 in 2020 to 16.3 in 2021, a 27.7% increase (Table). YLL per death did not change by more than 2.3% for any other cause (Figure).

**Figure.**
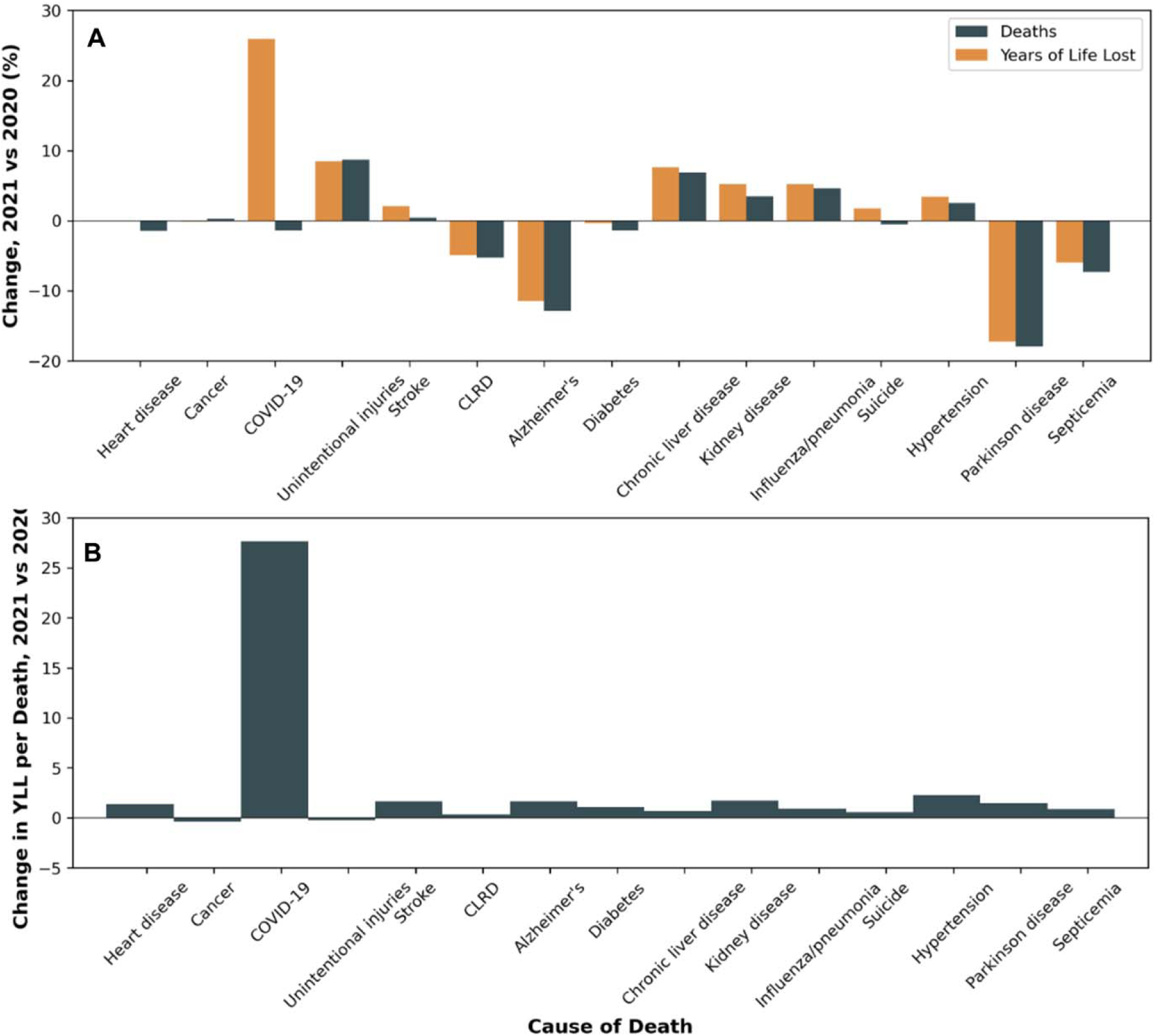
Changes in Leading Causes of Death and Associated Years of Life Lost, March to December 2020 and January to October 2021. For both panels, outcomes are compared between January through October 2021 and March through December 2020. Unintentional injury deaths include transportation incidents, unintentional drug overdoses and alcohol-related deaths, and other unintentional injury deaths. COVID-19 indicates coronavirus disease 2019; CLRD: chronic lower respiratory disease. **A**. Changes in Leadings Causes of Death and YLL. Paired bars indicate the percentage difference between study intervals for each outcome. **B**. Changes in YLL per Death. Bars indicate the percentage difference between study intervals for the outcome.

## 4. Discussion

We estimate that over the first 20 months of the pandemic, over 10 million years of life were lost directly due to COVID-19 deaths in the United States alone. Although COVID-19 deaths decreased slightly from 2020 to 2021, the number of YLL increased by 26% (over a million YLL). Increased YLL per COVID-19 death in 2021 result from younger-age COVID-19 mortality,^2^ contributing to more premature deaths caused by COVID-19 at a later stage of the pandemic despite advancements in vaccines in treatments. These findings underscore the need for sustained efforts to protect against COVID-19 mortality across age groups, especially given that this measure does not capture morbidity associated with post-acute sequelae of SARS-CoV-2 infection (i.e., Long COVID).

YLL from COVID-19 deaths in 2021 eclipsed YLL from unintentional injury deaths in 2020. Had unintentional injury deaths not increased by approximately 15,000 in 2021 as compared with 2020 (resulting in nearly 450,000 more YLL from unintentional injuries), COVID-19 deaths would have risen to the fourth-leading cause of YLL in 2020 to the third-leading cause of YLL in 2021.

Strengths of this analysis include the utilization of national datasets and sex- and age-specific standard life expectancies to estimate YLL. Limitations include provisional 2021 deaths, which are subject to reporting lags, and pre-pandemic standard life expectancies, as life expectancy in the US has declined. Importantly, the YLL metric compares an individual’s life expectancy with their age at the time of their death and should not be used as a measure of an individual’s potential contributions to society.

In conclusion, increased US COVID-19 deaths among younger people in the second year of the pandemic contributed to a marked increase in YLL from this preventable cause of death.

## Data Availability

All data produced are available online at CDC Wonder and the National Vital Statistics System.

https://wonder.cdc.gov/

https://www.cdc.gov/nchs/nvss/life-expectancy.htm

## Author Contributions

M.É.C. had full access to all the publicly available data in the study and takes responsibility for the integrity of the data and the accuracy of the data analysis.

## Concept and design

M.É.C.

## Drafting of the manuscript

M.É.C.

## Critical revision of the manuscript for important intellectual content

Both authors.

## Statistical analysis

M.É.C.

## Supervision

C.A.C.

## Financial Support

No direct funding supported this analysis. C.A.C. is the incumbent of an endowed professorship provided to Harvard University by Cephalon, Inc.

## Disclosures

M.É.C. received consultant payments from Vanda Pharmaceuticals and institutional grants paid to Monash University from the CDC and CDC Foundation, with funding provided by BNY Mellon, and from WHOOP, Inc., as well as an institutional gift to Monash University from Hopelab, Inc. C.A.C. reports grants/contracts to BWH from Dayzz Live Well, Delta Airlines, Jazz Pharma, Puget Sound Pilots, Regeneron Pharmaceuticals/Sanofi; is/was paid consultant/speaker for Institute of Digital Media and Child Development, Klarman Family Foundation, National Council for Mental Wellbeing, National Sleep Foundation, Physician’s Seal, SRS Foundation, Tencent, Teva Pharma Australia, With Deep, and Vanda Pharmaceuticals; holds an equity interest in With Deep and Vanda Pharmaceuticals; received travel support from Aspen Brain Institute, Bloomage International, Dr. Stanley Ho Medical Development Foundation, German National Academy of Sciences, National Safety Council, National Sleep Foundation, Stanford Medical School, and Vanda; receives research/education gifts through BWH from Arbor Pharmaceuticals, Avadel Pharmaceuticals, Bryte, Alexandra Drane, DR Capital Ltd, Eisai, Harmony Biosciences, Jazz Pharmaceuticals, Johnson and Johnson, Mary Ann & Stanley Snider via Combined Jewish Philanthropies, NeuroCare, Inc., Optum, Philips Respironics, Regeneron, Regional Home Care, ResMed, San Francisco Bar Pilots, Sanofi, Schneider, Simmons, Sleep Cycle. Sleep Number, Sysco, Teva Pharmaceuticals, Vanda; is/was an expert witness in legal cases, including those involving Advanced Power Technologies, Aegis Chemical Solutions, Amtrak; Casper Sleep Inc., C and J Energy Services, Catapult Energy Services Group, Covenant Testing Technologies, Dallas Police Association, Enterprise Rent-A-Car, Espinal Trucking/Eagle Transport Group/Steel Warehouse Inc., FedEx, Greyhound, PAR Electrical Contractors, Product and Logistics Services LLC/Schlumberger Technology, Puckett EMS, Puget Sound Pilots, Union Pacific Railroad, UPS, and Vanda; royalties from Philips Respironics for the Actiwatch-2 and Actiwatch Spectrum devices. C.A.C.’s interests were reviewed and are managed by the BWH and MGB in accordance with their conflict-of-interest policies. No other disclosures were reported.

## Notes

### Author Declarations

The study used openly available human data that were originally located at CDC WONDER and the National Vital Statistics System.

